# Developing and optimizing machine learning algorithms for predicting in-hospital patient charges for Congestive Heart Failure Exacerbations, Chronic Obstructive Pulmonary Disease Exacerbations and Diabetic Ketoacidosis

**DOI:** 10.1101/2023.12.17.23298944

**Authors:** Monique Arnold, Lathan Liou, Mary Regina Boland

## Abstract

**Background:** Hospitalizations for exacerbations of congestive heart failure (CHF), chronic obstructive pulmonary disease (COPD) and diabetic ketoacidosis (DKA) are costly in the United States.

**Objective:** The purpose of this study is to predict in-hospital charges for each condition using Machine Learning (ML) models.

**Methods:** We conducted a retrospective cohort study on national discharge records of hospitalized adult patients from January 1^st^, 2016, to December 31^st^, 2019. We used numerous ML techniques to predict in-hospital total cost.

**Results:** We found that linear regression (LM), gradient boosting (GBM) and extreme gradient boosting (XGB) models had good predictive performance and were statistically equivalent, with training R-Squared values ranging from 0.49–0.95 for CHF; 0.56–0.95 for COPD; and 0.32–0.99 for DKA. We identified important key features driving costs, including patient age, length-of-stay, number of procedures. and elective/non-elective admission.

**Conclusions:** ML methods may be used to accurately predict costs and identify drivers of high cost for COPD exacerbations, CHF exacerbations and DKA. Overall, our findings may inform future studies that seek to decrease the underlying high patient costs for these conditions.

## INTRODUCTION

### 1.1 Background on Healthcare Costs Associated with Our Outcomes: CHF, COPD, DKA

In the United States, hospital expenditures account for approximately one-third of overall healthcare expenditures with an estimated total of US$1.192 billion in 2019 according to the Center for Medicare & Medicaid Services.^1^Healthcare costs are disproportionately concentrated among a small group of high-cost patients.^2–4^ High-cost patients often have significant unmet critical healthcare needs despite the substantial healthcare costs they incur.^5,6^

Congestive heart failure (CHF), chronic obstructive pulmonary disease (COPD) and diabetes mellitus, are life-altering, high-cost, high-volume conditions that affect millions of people and result in many hospitalizations per year.^7^ According to Medical Expenditure Panel Survey data for 2017 to 2018 published by the American Heart Association (AHA), diabetes mellitus, heart disease including CHF and respiratory conditions including COPD were in the top 10 leading diagnoses for direct health expenditures.^8^

CHF is one of the leading causes of hospitalization in the U.S., affecting 6 million adults as of 2018 and costing the nation an estimated $30.7 billion in 2012 according to the American Heart Association, with these costs deriving largely from exacerbations requiring emergency visits and hospitalizations.^8–10^ Similarly, COPD is a high-cost disease–as COPD progresses, patients often experience acute exacerbations, characterized by dyspnea, cough, sputum production and worsening lung function; COPD exacerbations cause frequent hospital admissions and readmissions, reportedly accounting for 90.3% of the total medical cost related to COPD and leading to US $32.1 billion in total medical cost.^11,12^Finally, diabetic ketoacidosis (DKA) is one of the acute, life-threatening complications of diabetes mellitus, a disease affecting 37.3 million people as of 2019 according to the CDC.^13^ DKA is a common cause of hospitalization in patients with diabetes, and is characterized by uncontrolled hyperglycemia, metabolic acidosis, and increased serum ketone concentration.^14,15^

### 1.2 Prior Machine Learning Methods Studying Our Outcomes: CHF, COPD, DKA

Machine Learning (ML) techniques have emerged as a mechanism to analyze high-dimensional medical data to understand factors underlying patient, hospital-level and health system-level outcomes.^16^ Specifically, for our three cohorts of patients, ML techniques have been utilized to identify at-risk patients, predict risk of readmissions and readmission rates, and to predict length of inpatient stay.^11,12,17–21^ Work has been done to develop predictive models to identify major underlying drivers of high healthcare costs for patients in generalized cohorts as well as several other cohort of patients, for example, breast cancer patients and coronary artery bypass graft patients.^22–26^ To date, however, robust machine learning algorithms for predicting in-hospital expenditures and the factors that influence them have not been evaluated in patients experiencing CHF exacerbations, COPD exacerbations or DKA.

## OBJECTIVES

The purpose of our study is to build and evaluate ML models to predict in-hospital charges associated with hospitalizations for these conditions, as this has not been done previously. Furthermore, based on the model output, we provide recommendations for model optimality in modeling in-hospital expenditures in each cohort, as well as identify factors that underlie high-cost in-hospital admissions for each of the three diseases.

## METHODS

An overview of the methodology employed is shown in **Figure 1**. All data processing and statistical and machine learning analyses were conducted using R version 4.1.1 (version “Kick Things”, released August 8, 2021) and RStudio Version 1.4.1717.

**Figure. 1.**
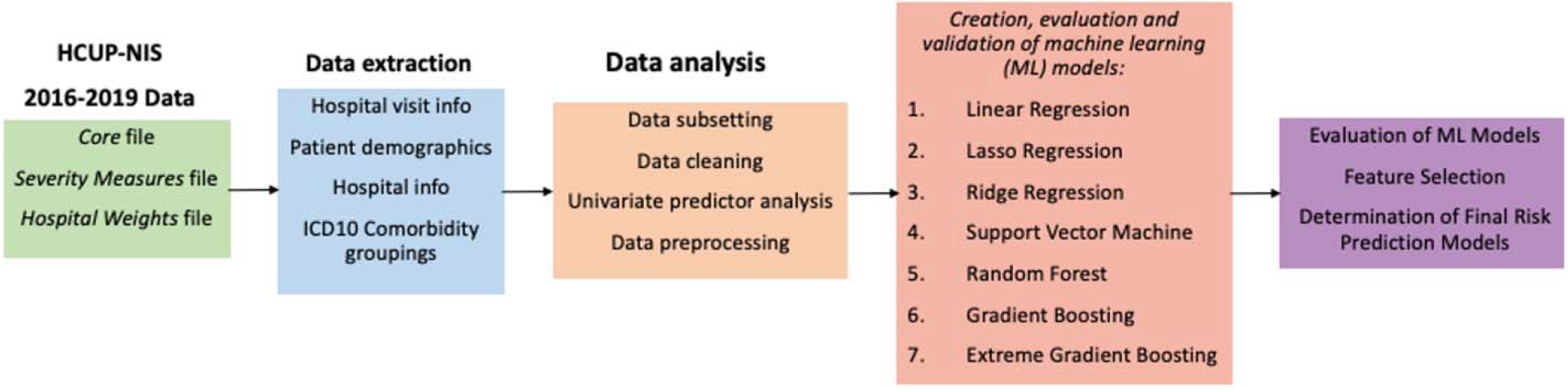
Overview of study. The HCUP-NIS 2016 Core, Severity Measures, Hospital Weights, and Cost Charge files were merged, and data related to hospital discharges and demographics were extracted as continuous, categorical and binary variables. ICD-10 comorbidity mappings from AHRQ were determined from ICD-10 codes. R codes were written to extract, clean and analyze the HCUP-NIS data. Seven ML models were then trained, evaluated, and validated for each of the three disease cohorts, and the best model for each disease cohort was determined.

### 3.1 Dataset and Study Design

The National (Nationwide) Inpatient Sample (NIS) is a large, publicly available all-payer inpatient care database in the United States, containing data on more than seven million hospital discharges each year, and is maintained as part of the Healthcare Cost and Utilization Project (HCUP).^27–29^ We used the HCUP-NIS Core, Severity, Hospital and Cost Charge datasets and queried the datasets for all hospitalizations between January 1, 2016 and December 31, 2019. Hospital discharges in which the patient was aged < 18 years or died were excluded.

We identified patients matching the three disease conditions using the International Classification of Diseases version 10 (ICD-10) codes: 1) Chronic Obstructive Pulmonary Disease (COPD) exacerbations via ICD-10 code J441; 2) Congestive Heart Failure (CHF) exacerbations via ICD-10 codes I5021, I5023, I5031, I5033, I5041, I5043; 3) Diabetic ketoacidosis without coma (DKA) via ICD-10 codes E1010, E1011, E1111, E1110.^30^ **Supplemental Table 1** shows the extracted ICD-10 codes and principal diagnoses for each of these conditions.

We identified a total of 26,190 unique discharges across the three conditions, including 9,552 discharges for COPD, 14,688 for CHF and 1,950 discharges for DKA. The primary outcome for this study was total hospital charges (totchg in the HCUP-NIS Core datasets). This cohort was identified after excluding discharges with missing data for any of the predictor variables of interest (as further described below).

### 3.2 Predictor variables

We conducted a preliminary literature review to determine potential factors that may affect in-hospital charges and that could be used as predictors in our analysis. Initial predictors for analysis included 46 variables, including 29 unique ICD-10 diagnosis code groupings, extracted from HCUP-NIS dataset, which included demographic characteristics, hospital-related variables, health care utilization six months before index admission, and discharge-related variables. A brief description for each predictor variable is given in **Supplemental Table 2**. Further description of the potential values of each variable can be found on the NIS Description of Data Elements page (https://www.hcup-us.ahrq.gov/db/nation/nis/nisdde.jsp).

ICD10 diagnosis codes were transformed into Agency for Healthcare Research and Quality (AHRQ) comorbidity categories using the icd R package. If a patient had at least one ICD10 code in one of the AHRQ comorbidity categories, then they were considered positive for that category. A list of AHRQ comorbidity categories is shown in **Supplemental Table 3**.

### 3.3 Univariable analysis of predictor variables

The relationships between each of the predictor variables and total charges were analyzed using two-sample t-tests. Statistical significance was determined at the 95% confidence level, with p < 0.05 noted as statistically significant. We also calculated the correlations between each predictor variable in the dataset using the Pearson method. To reduce the sheer quantity of variables without having to choose variables a priori, only variables with a Pearson correlation coefficient above 0.2 were visualized.

### 3.4 Preprocessing of variables

Due to the asymmetric distribution of characteristics and predictor variables, cases with missing data for any of the dependent or independent variables were excluded from this analysis, a common, though controversial, approach for dealing with missing values.^31^ For the ML analysis, “one-hot encoding” was performed, in which each categorical variable was transformed into a numerical dummy variable.^32^ With one-hot encoding, a total of 79 predictor variables were used. Additionally, all continuous or numerical variables, including total charges, were standardized such that their mean was 0 and standard deviation was 1. This is a common preprocessing method used to decrease the likelihood of bias of the model due to very large or small numeric variables.^33^

### 3.5 ML models

We used seven ML algorithms, namely linear regression (LM), lasso regression (LASSO), ridge regression (RIDGE), support vector machine (SVM), random forest (RF), gradient boosting (GBM) and extreme gradient boosting (XGB). These have been previously used in healthcare machine learning to build models for healthcare classification and prediction models. Models were trained and tested using the caret package in R. For training and validation of the model, a five-fold cross-validation using 75% of the derivation sample for development with validation at 25% was conducted. The ML models were developed on the training set and then validated on the testing set.

The five-fold cross-validation approach was used to obtain reliable results for evaluating prediction models or for obtaining reliable results. Specifically, the original training set was split into five folds through stratified random sampling. For the *i*th iteration, fold *i* was treated as the validation set and the remaining four folds were used to train the model. The procedure was repeated for five iterations. This process allows for the model performance to be estimated over all the data.

### 3.6 Model Evaluation and Comparison

Models were evaluated based on root-mean square error (RMSE) and R-squared values of the models, common metrics used to measure the accuracy of prediction models.^34,35^ RMSE measures the quality of predictions by determining how far predictions fall from measured true values using Euclidean distance. It is a standard metric to measure the error of a model, with smaller values indicating less random noise and thus higher accuracy. R-squared is a measure goodness of fit of a model and has a maximum value of 1. Models with R-squared values closer to 1 are more well fitted to the data. We compared models using paired samples t-tests to determine if the differences between them were statistically significantly different at the 95% confidence level.

### 3.7 Feature Selection

We performed feature selection in two ways. First, relative importance of predictor variables (i.e., feature importance) were determined from the ML models and reported as variable importance (VI) scores. VI scores demonstrate how much the prediction changes as the feature values vary.^36^ Higher feature importance indicates the higher importance of the feature to the model prediction. Documentation for the caret package indicates that for linear models, the relative importance is determined by the absolute value of the t-statistic. For gradient boosting models, the relative importance is determined from the absolute value of the coefficients corresponding to the tuned model. All importance values were scaled from 0 to 100. Based on this relative feature importance, we visualized the top twenty most influential features.

Secondly, using the caret package, we performed recursive feature elimination (RFE), which employs backwards selection algorithms to determine the most important features for prediction in each condition using linear functions **(Supplementary Figure 2)**.^37^ First, the algorithm fits the model to all predictors and each predictor is ranked using its importance to the model. From *i* equals 1 to 50, model is iterated with *i* number of features, and at each iteration of feature selection, the *i* top ranked predictors are retained, then the model is refit and performance is assessed. The value of *i* with the best performance is determined and the top *i* predictors are then used to fit the final models.

### 3.8 Ethical Considerations

We obtained Institutional Review Board approval of this study from the University of Pennsylvania (protocol #851472).

## RESULTS

### 4.1 Sample characteristics

In total, 26,190 unique hospital discharges with complete data were available for the analysis from January 1, 2016, to December 31, 2019–14,688 patients hospitalized for a CHF exacerbation, 9,552 patients hospitalized for COPD exacerbation and 1,950 patients hospitalized for DKA without coma. Characteristics of the sample cohorts are summarized in **Table 1**. Average costs for hospitalizations were US$18,196 (± $29,248) for CHF exacerbations, US$13,572 (± $17,598) for COPD exacerbations and $13,650 (± $16,778) for DKA episodes. Mean length of stay and number of inpatient procedures were highest in the CHF cohort at 6.36 days and 1.90 procedures respectively; mean length of stay was 5.32 days in the COPD exacerbation cohort and 5.08 in the DKA cohort, and number of procedures was 1.32 each for COPD patients and DKA patients. As shown in **Figure 2**, the mean cost charges for each condition steadily increased for each condition over the four-year period from 2016 to 2019.

**Table 1.** Overall Patient Cohort Demographics and Characteristics for Each Disease: CHF exacerbations, COPD exacerbations ad DKA episodes.

|  | CHF<br>Exacerbation | COPD<br>Exacerbation | DKA<br>Episode | Overall |
| --- | --- | --- | --- | --- |
| **n** | 14688 | 9552 | 1950 | 26190 |
| **Total Charges (\$, mean (SD))** | 18196.21<br>(29247.63) | 13572.25<br>(17598.13) | 13650.48<br>(16777.65) | 16171.31<br>(24876.85) |
| **Length of Stay in days (mean (SD))** | 6.36 (7.26) | 5.32 (5.74) | 5.08 (6.75) | 5.89 (6.73) |
| **Number of Procedures (mean (SD))** | 1.90 (2.82) | 1.32 (2.13) | 1.32 (2.31) | 1.64 (2.57) |
| **Elective Admission = Yes (%)** | 1910 (13.0) | 1155 (12.1) | 194 (9.9) | 3259 (12.4) |
| **Sex = Female (%)** | 7521 (51.2) | 5385 (56.4) | 1029 (52.8) | 13935 (53.2) |
| **Age (mean (SD))** | 66.77 (17.84) | 64.92 (16.51) | 50.11 (19.78) | 64.85 (18.03) |
| **Race (%)** |  |  |  |  |
| White | 10500 (71.5) | 7243 (75.8) | 1208 (61.9) | 18951 (72.4) |
| Black | 2166 (14.7) | 1165 (12.2) | 402 (20.6) | 3733 (14.3) |
| Hispanic | 1197 (8.1) | 663 (6.9) | 214 (11.0) | 2074 (7.9) |
| Asian Pacific Islander | 374 (2.5) | 195 (2.0) | 54 (2.8) | 623 (2.4) |
| Native American | 73 (0.5) | 56 (0.6) | 23 (1.2) | 152 (0.6) |
| Other | 378 (2.6) | 230 (2.4) | 49 (2.5) | 657 (2.5) |
| **Insurance status (%)** |  |  |  |  |
| Medicare | 9621 (65.5) | 6086 (63.7) | 707 (36.3) | 16414 (62.7) |
| Medicaid | 1827 (12.4) | 1345 (14.1) | 504 (25.8) | 3676 (14.0) |
| PrivateInsurance | 2570 (17.5) | 1647 (17.2) | 514 (26.4) | 4731 (18.1) |
| SelfPay | 375 (2.6) | 268 (2.8) | 162 (8.3) | 805 (3.1) |
| NoCharge | 23 (0.2) | 22 (0.2) | 10 (0.5) | 55 (0.2) |
| Other | 272 (1.9) | 184 (1.9) | 53 (2.7) | 509 (1.9) |
| **Median household income quartile for patient ZIP Code (%)** |  |  |  |  |
| 0 to 25th percentile | 4017 (27.3) | 2877 (30.1) | 635 (32.6) | 7529 (28.7) |
| 26th to 50th percentile | 3771 (25.7) | 2613 (27.4) | 498 (25.5) | 6882 (26.3) |
| 51st to 75th percentile | 3767 (25.6) | 2314 (24.2) | 464 (23.8) | 6545 (25.0) |
| 76th to 100th percentile | 3133 (21.3) | 1748 (18.3) | 353 (18.1) | 5234 (20.0) |
| **Discharge (%)** |  |  |  |  |
| Routine | 7633 (52.0) | 5545 (58.1) | 1393 (71.4) | 14571 (55.6) |
| Transfer to Hospital | 321 (2.2) | 175 (1.8) | 38 (1.9) | 534 (2.0) |
| Transfer to Other Facility | 3578 (24.4) | 1917 (20.1) | 278 (14.3) | 5773 (22.0) |
| Home Health Care | 3155 (21.5) | 1914 (20.0) | 241 (12.4) | 5310 (20.3) |
| Unknown | 1 (0.0) | 1 (0.0) | 0 (0.0) | 2 (0.0) |
| **Patient Location (%)** |  |  |  |  |
| "Central" counties of metro areas of >=1 million population | 4421 (30.1) | 2712 (28.4) | 626 (32.1) | 7759 (29.6) |
| "Fringe" counties of metro areas of >=1 million population | 3688 (25.1) | 2404 (25.2) | 449 (23.0) | 6541 (25.0) |
| Counties in metro areas of 250,000-999,999 population | 2953 (20.1) | 1876 (19.6) | 408 (20.9) | 5237 (20.0) |
| Counties in metro areas of 50,000-249,999 population | 1361 (9.3) | 981 (10.3) | 195 (10.0) | 2537 (9.7) |
| Micropolitan counties | 1160 (7.9) | 811 (8.5) | 141 (7.2) | 2112 (8.1) |
| Not metropolitan or micropolitan counties | 1105 (7.5) | 768 (8.0) | 131 (6.7) | 2004 (7.7) |
| **Hospital Division (%)** |  |  |  |  |
| NewEngland | 1190 (8.1) | 546 (5.7) | 92 (4.7) | 1828 (7.0) |
| MiddleAtlantic | 3094 (21.1) | 2100 (22.0) | 368 (18.9) | 5562 (21.2) |
| EastNorthCentral | 866 (5.9) | 547 (5.7) | 79 (4.1) | 1492 (5.7) |
| WestNorthCentral | 3581 (24.4) | 2480 (26.0) | 493 (25.3) | 6554 (25.0) |
| SouthAtlantic | 1370 (9.3) | 871 (9.1) | 166 (8.5) | 2407 (9.2) |
| EastSouthCentral | 1240 (8.4) | 942 (9.9) | 165 (8.5) | 2347 (9.0) |
| WestSouthCentral | 716 (4.9) | 375 (3.9) | 130 (6.7) | 1221 (4.7) |
| Mountain | 522 (3.6) | 377 (3.9) | 102 (5.2) | 1001 (3.8) |
| Pacific | 2109 (14.4) | 1314 (13.8) | 355 (18.2) | 3778 (14.4) |
| **Hospital Bedsize (%)** |  |  |  |  |
| Small | 2044 (13.9) | 1621 (17.0) | 278 (14.3) | 3943 (15.1) |
| Medium | 3497 (23.8) | 2526 (26.4) | 467 (23.9) | 6490 (24.8) |
| Large | 9147 (62.3) | 5405 (56.6) | 1205 (61.8) | 15757 (60.2) |
| **Hospital Location/Teaching Status (%)** |  |  |  |  |
| Rural | 916 (6.2) | 808 (8.5) | 126 (6.5) | 1850 (7.1) |
| UrbanNonTeaching | 2431 (16.6) | 1931 (20.2) | 323 (16.6) | 4685 (17.9) |
| UrbanTeaching | 11341 (77.2) | 6813 (71.3) | 1501 (77.0) | 19655 (75.0) |
| **Hospital Control/Ownership (%)** |  |  |  |  |
| Government, nonfederal | 1653 (11.3) | 1101 (11.5) | 295 (15.1) | 3049 (11.6) |
| Private, not-profit | 11710 (79.7) | 7396 (77.4) | 1464 (75.1) | 20570 (78.5) |
| Private, invest-own | 1325 (9.0) | 1055 (11.0) | 191 (9.8) | 2571 (9.8) |

**Figure 2.**
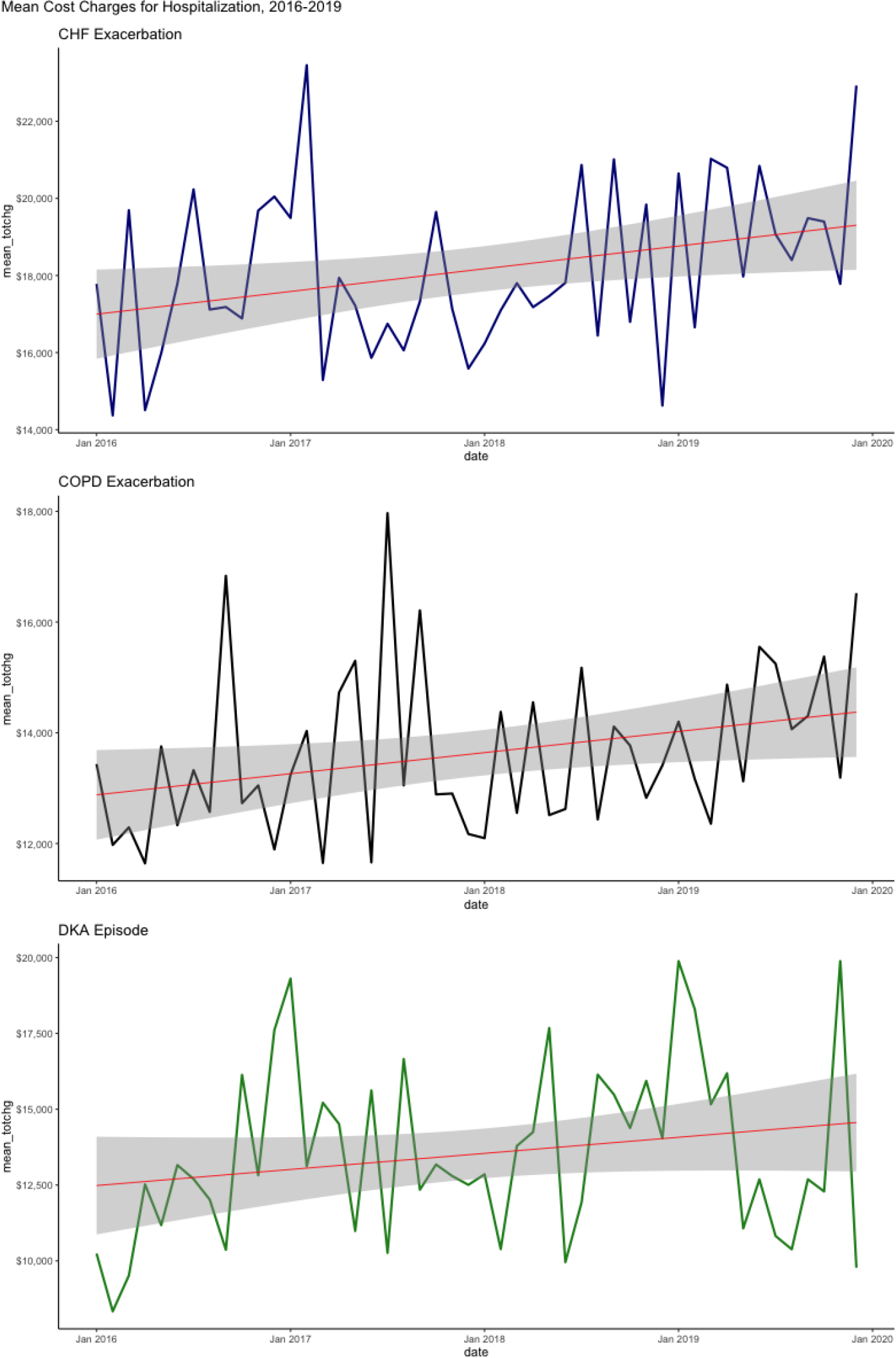
Mean cost charges. Trends in mean cost charges for hospitalization for each condition, 2016-2019.

### 4.2 Univariable analyses

**Tables 2** and **Table 3** show the univariable results for the categorical and continuous variables, respectively. Longer inpatient stay and higher number of procedures were associated with higher in-hospital total charges. Older patients also incurred higher total charges. For several features of sex, payment method, hospital bedsize, hospital control, hospital location, All Patients Refined Diagnosis Related Groups (APRDRG) severity score and APRDRG risk mortality score, the difference in total charges between groups of patients within each cohort were often statistically significant (for example, patients in large hospitals incurred greater charges than those in smaller hospitals in each disease cohorts, p < 0.05). Notably, Black patients incurred higher charges than White patients (p < 0.01).

**Table 2.** Univariable results for the categorical variables.

|  | CHF |  |  | COPD |  |  | DKA |  |  |
| --- | --- | --- | --- | --- | --- | --- | --- | --- | --- |
|  | Mean Charges | p-value |  | Mean Charges | p-value |  | Mean Charges | p-value |  |
| **Sex** |  |  |  |  |  |  |  |  |  |
| Male | \$20,362 | | | \$14,909 | | | \$14,095 | | |
| Female | \$16,132 | 0.00 | **** | \$12,538 | 0.00 | **** | \$13,253 | 0.27 | ns |
| **Race** |  |  |  |  |  |  |  |  |  |
| White | \$18,067 | | | \$13,319 | | | \$13,926 | | |
| Black | \$17,379 | 0.60 | ns | \$13,645 | 0.84 | ns | \$10,917 | 0.01 | ** |
| Hispanic | \$18,139 | 0.94 | ns | \$13,812 | 0.84 | ns | \$15,967 | 0.48 | ns |
| AsianPacificIslander | \$21,378 | 0.24 | ns | \$17,079 | 0.16 | ns | \$14,833 | 0.84 | ns |
| NativeAmerican | \$18,276 | 0.94 | ns | \$14,366 | 0.84 | ns | \$18,703 | 0.60 | ns |
| Other | \$23,489 | 0.16 | ns | \$17,326 | 0.16 | ns | \$15,487 | 0.84 | ns |
| **Payment Method** |  |  |  |  |  |  |  |  |  |
| PrivateInsurance | \$19,193 | | | \$13,285 | | | \$12,951 | | |
| Medicare | \$18,070 | 0.35 | ns | \$13,758 | 0.48 | ns | \$16,398 | 0.00 | ** |
| Medicaid | \$17,609 | 0.31 | ns | \$13,264 | 0.98 | ns | \$12,071 | 0.48 | ns |
| SelfPay | \$15,916 | 0.11 | ns | \$11,673 | 0.35 | ns | \$9,106 | 0.00 | ** |
| NoCharge | \$9,612 | 0.00 | ** | \$11,102 | 0.48 | ns | \$5,410 | 0.00 | ** |
| Other | \$21,041 | 0.42 | ns | \$15,327 | 0.21 | ns | \$14,246 | 0.64 | ns |
| **Hospital Bedsize** |  |  |  |  |  |  |  |  |  |
| Small | \$13,775 | | | \$12,446 | | | \$12,447 | | |
| Medium | \$15,270 | 0.00 | ** | \$12,862 | 0.41 | ns | \$11,750 | 0.51 | ns |
| Large | \$20,303 | 0.00 | **** | \$14,242 | 0.00 | **** | \$14,665 | 0.04 | * |
| **Hospital Location** |  |  |  |  |  |  |  |  |  |
| Rural | \$11,894 | | | \$11,007 | | | \$10,736 | | |
| UrbanNonTeaching | \$13,773 | 0.00 | ** | \$11,706 | 0.13 | ns | \$11,896 | 0.30 | ns |
| UrbanTeaching | \$19,653 | 0.00 | **** | \$14,405 | 0.00 | **** | \$14,273 | 0.00 | *** |
| **Hospital Control** |  |  |  |  |  |  |  |  |  |
| Government | \$18,839 | | | \$14,660 | | | \$14,077 | | |
| PrivateNotProfit | \$18,593 | 0.87 | ns | \$13,649 | 0.29 | ns | \$13,905 | 0.87 | ns |
| PrivateInvestOwn | \$13,887 | 0.00 | **** | \$11,896 | 0.00 | ** | \$11,043 | 0.06 | ns |
| **APRDRG Severity** |  |  |  |  |  |  |  |  |  |
| MinorLOF | \$16,181 | | | \$11,425 | | | \$10,375 | | |
| ModerateLOF | \$16,092 | 0.88 | ns | \$11,796 | 0.38 | ns | \$11,615 | 0.15 | ns |
| MajorLOF | \$18,092 | 0.00 | ** | \$14,015 | 0.00 | **** | \$14,894 | 0.00 | **** |
| ExtremeLOF | \$29,323 | 0.00 | **** | \$23,751 | 0.00 | **** | \$26,027 | 0.00 | **** |
| NoClass | \$21,596 | 0.65 | ns | \$10,116 | 0.80 | ns | | | |
| **APRDRG Risk Mortality** |  |  |  |  |  |  |  |  |  |
| MinorLklhdDying | \$16,550 | | | \$11,534 | | | \$10,724 | | |
| ModerateLklhdDying | \$16,349 | 0.72 | ns | \$12,733 | 0.00 | *** | \$13,435 | 0.00 | ** |
| MajorLklhdDying | \$18,513 | 0.00 | *** | \$14,786 | 0.00 | **** | \$16,753 | 0.00 | **** |
| ExtremeLklhdDying | \$28,668 | 0.00 | **** | \$23,321 | 0.00 | **** | \$23,799 | 0.00 | **** |
| NoClass | \$21,596 | 0.68 | ns | \$10,116 | 0.72 | ns | | | |

**Table 3.** Univariable results for the continuous variables.

|  | CHF |  |  |  | COPD |  |  |  | DKA |  |  |  |
| --- | --- | --- | --- | --- | --- | --- | --- | --- | --- | --- | --- | --- |
|  | Estimate | Std. Error | t value | Pr(>\|t\|) | Estimate | Std. Error | t value | Pr(>\|t\|) | Estimate | Std. Error | t value | Pr(>\|t\|) |
| Age | -1.38 | 10.90 | 4.16 | 0.00 | 45.36 | 10.90 | 4.16 | 0.00 | 173.85 | 18.81 | 9.24 | 0.00 |
| Length of Stay | 3024.61 | 21.94 | 137.86 | 0.00 | 2344.85 | 20.21 | 116.02 | 0.00 | 1694.71 | 41.16 | 41.17 | 0.00 |
| Number of Procedures | 6399.13 | 67.30 | 95.09 | 0.00 | 4880.51 | 68.06 | 71.71 | 0.00 | 4556.15 | 128.55 | 35.44 | 0.00 |

**Table 4.**
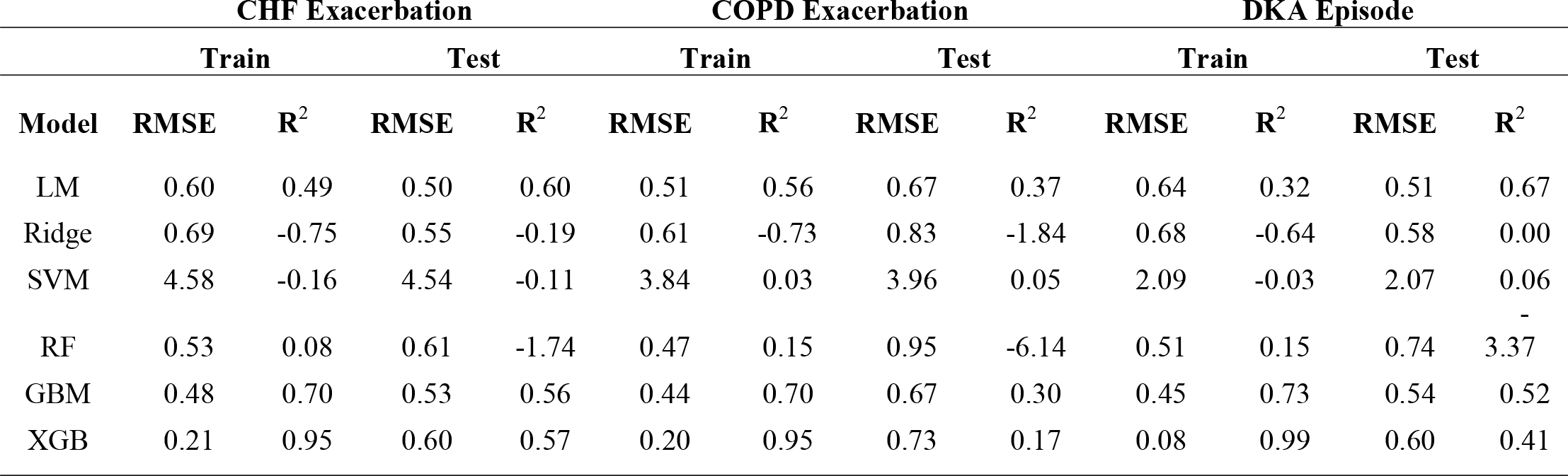
Comparison of metrics of ML models.

The Pearson correlation coefficients of the most correlated variables are visualized in **Figure 3**. The data show that collinearity exists between several variables. For each of the three conditions, number of procedures and APRDRG risk mortality were the most strongly positively correlated of the non-diagnosis variables (with correlation coefficients of 0.80 for CHF, 0.79 for COPD and 0.77 for DKA) while age and payment method were the most negatively correlated of the non-diagnosis variables (with correlation coefficients of -0.50 for CHF, -0.50 for COPD and -0.44 for DKA).

**Figure 3.**
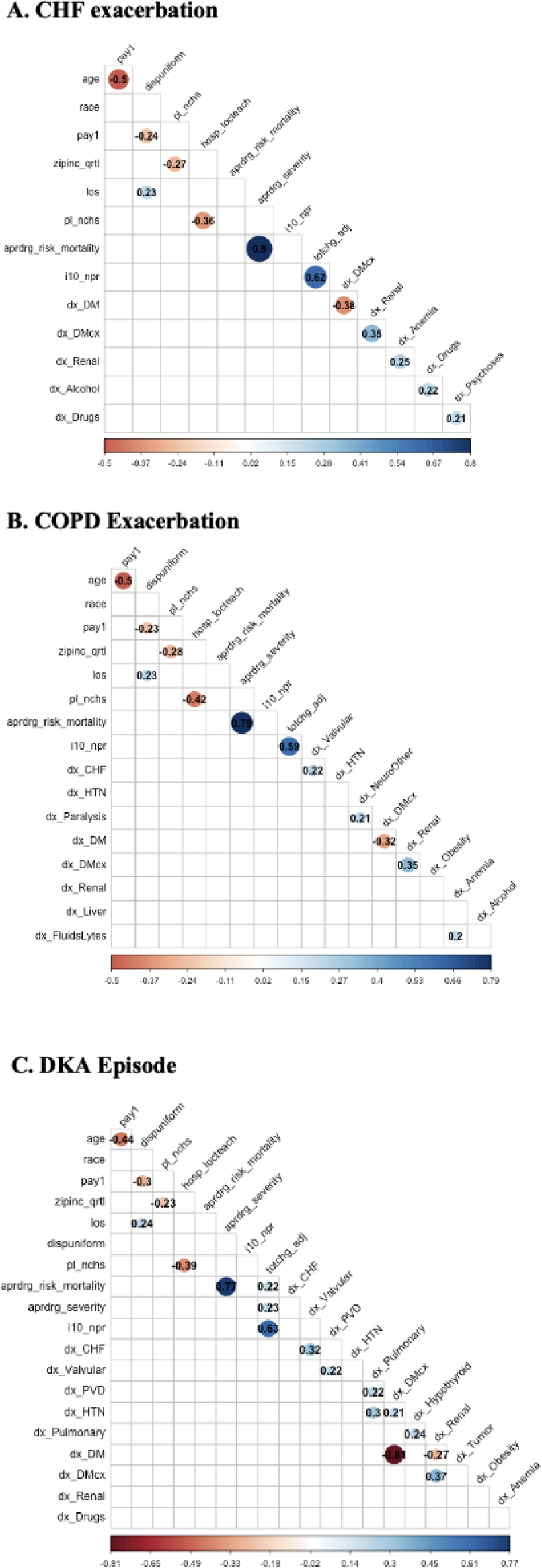
Correlation plots for each disease condition. Only those variables with a Pearson coefficient > 0.2 are displayed. Pearson correlation coefficients are displayed.4.3 Model Evaluation and Comparison Accuracy metrics for the machine learning models are shown in Table 4. Among the 6 ML algorithms, the LM, GBM and XGB models had the best performances across the three conditions.

**Figure 4** shows boxplots of the accuracy metrics for the out-of-sample performance within each “sample” for these three models. Pairwise samples t-tests showed that the differences between each of the three models for each condition were not statistically significant at the 95% confidence level and, as such, within each disease condition the LM, GBM and XGB models were equivalent. RMSEs for the training model ranged from 0.21 to 0.60 and R-Squared from 0.49 to 0.95 for CHF; RMSEs ranged from 0.20 to 0.51 and R-Squared from 0.56 to 0.95 for COPD; and RMSEs ranged from 0.08 to 0.64 and R-Squared from 0.32 to 0.99 for DKA. RMSEs for the test model ranged from 0.50 to 0.60 and R-Squared from 0.56 to 0.60 for CHF; RMSEs ranged from 0.67 to 0.73 and R-Squared from 0.17 to 0.37 for COPD; and RMSEs ranged from 0.51 to 0.60 and R-Squared from 0.41 to 0.67 for DKA.

**Figure 4.**
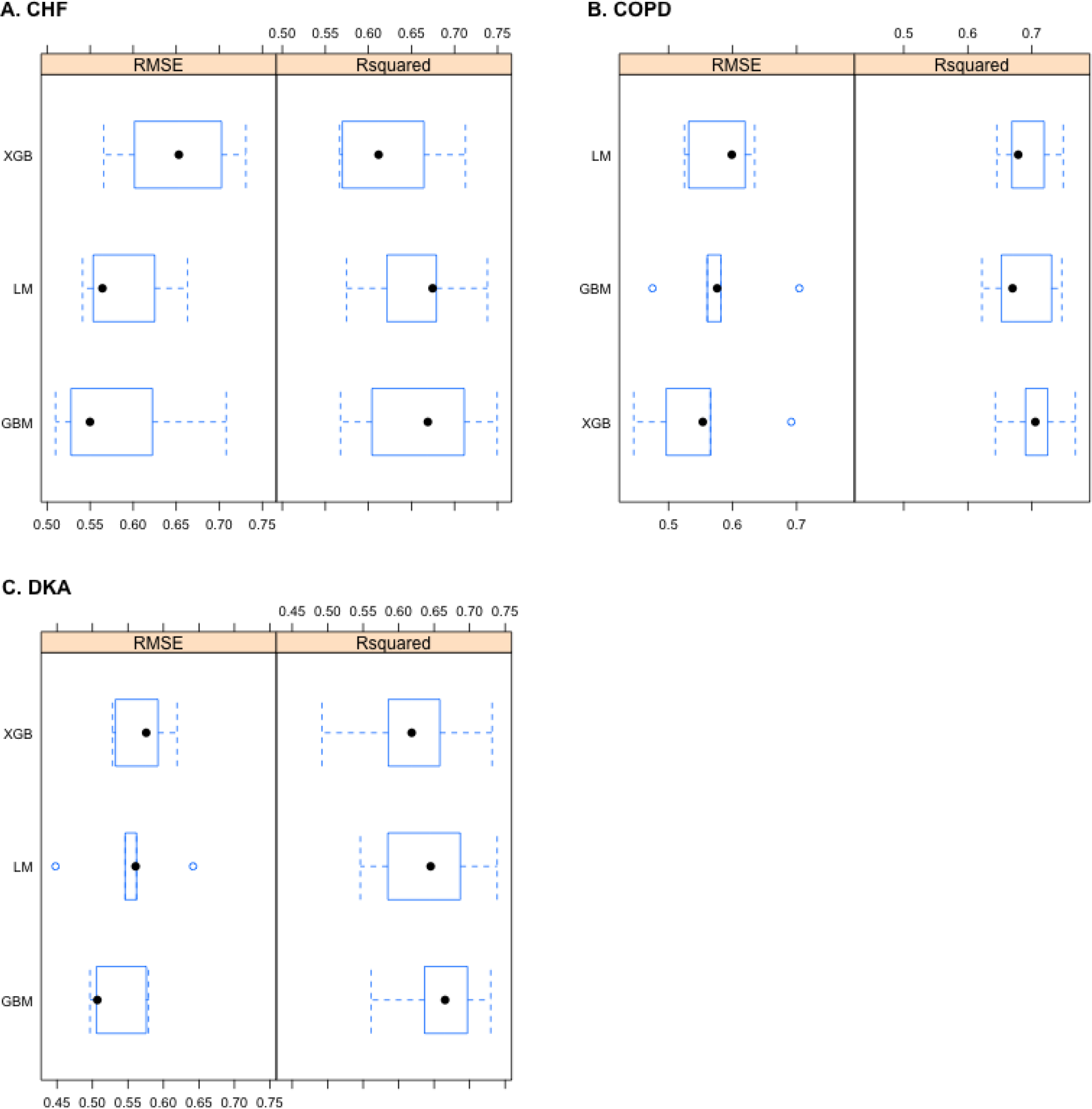
Comparison of models. Comparison of RMSE and R-squared for the out-of-sample performance within each resample for the LM, GBM and XGB models for each disease condition

### 4.4 Feature Selection

Top 20 features in each model determined from the training LM, GBM and XGB models for each condition were determined **(Supplemental Figure 1)**. Length of stay was the most important predictor in each of the models, followed by the number of procedures during hospitalization. Age and elective/non-elective admission were also important predictors in at least one model for each disease condition, but with much smaller VI scores than length of stay and age. This aligns with our univariable analyses **(Tables 2 and 3)**.

## DISCUSSION

Although many studies have employed ML techniques to predict at-risk patients, readmission risks, readmission rates and length of stay for CHF, COPD and DKA patients, the development of a predictive model of in-hospital cost charges in these disease cohorts is a novel contribution of this study.

We constructed 6 ML models for each disease and found that LM, GBM and XGB models performed the best–they had good predictive performance and were found to be statistically equivalent. Thus, traditional linear regression was not inferior to the tree-based models. Training metrics showed RMSEs ranged from 0.21–0.60 and R-Squared from 0.49–0.95 for CHF; RMSEs from 0.20–0.51 and R-Squared from 0.56–0.95 for COPD; and RMSEs 0.08–0.64 and R-Squared from 0.32–0.99 for DKA. The corresponding metrics for the test models were all lower than their training models, indicating the models performed worse on the validation datasets.

Unsurprisingly, length of stay was the most important predictor in each of the models, disproportionately affecting hospital charges in each model. This was followed by the number of procedures during hospitalization. Age and elective/non-elective admission were also important predictors in at least one model for each disease condition. Feature selection indicates that though these variables are extremely influential in any model, many other patient-level and hospital-level features also have small but measurable impacts on hospital charges.

### 5.1 Strengths of Our Study

The strengths of our study include the large sample size that the HCUP NIS datasets provide. Furthermore, the availability of many demographic characteristics, diagnosis-related variables, and hospital characteristics for use as predictors allowed for the building of supervised prediction models. The use of advanced ML techniques represents a robust use of data science to characterize a complex clinical issue. The ability to predict expenditures at the patient-level with good accuracy can allow for targeted care by anticipating health care needs of patients. This will provide insights into designing effective and tailored interventions to meet the needs of high-cost patients and reduce costs.

### 5.2 Limitations of Our Study

Despite its strengths, we recognize that this work has several limitations. Missing data is a well-known limitation of utilizing EMR data for research, of which the HCUP-NIS is susceptible. Additionally, we chose to use only complete data without missing values for all predictor variables, thereby losing a substantial amount of possible discharge events. Future work can involve employing data imputation methods rather than data exclusion. This could help to address the potential selection bias that can result from categorically excluding cases with missing data.

Additionally, the discharge data used may include discharges from readmissions of the same patient. The NIS data contains discharge-level records which, per the HCUP-NIS documentation, means that “individual patients who are hospitalized multiple times in one year may be present in the NIS multiple times… this will be especially important to remember for certain conditions for which patients may be hospitalized multiple times in a single year.”^29,38^ As discussed, our target patients often experience numerous hospitalizations, and initial versus recurrent hospitalizations might differ in their character. As such, we considered limiting the analysis to initial discharge, however “…there is no uniform patient identifier available that allows a patient-level analysis with the NIS.” Therefore, for the purposes of this study, we included all the discharges and performed the analysis at the discharge-level.

## CONCLUSION

We demonstrated the use of ML models to predict in-hospital charges for patients hospitalized for CHF exacerbation, COPD exacerbation and DKA. We found that length of stay, number of procedures during hospitalization, age and elective/non-elective admission were important predictors in these models for these diseases. This research can provide helpful information for medical management, which may decrease health insurance burdens in the future.

## Data Availability

All data produced in the present study are available upon reasonable request to the authors.

## ACKNOWLEDGEMENTS

Special thanks to the enrolled class members of University of Pennsylvania Spring 2022 BMIN 505: Precision Medicine and Healthy Policy for their insights and feedback.

## SUPPLEMENTARY FILES

**Supplemental Figure 1.**
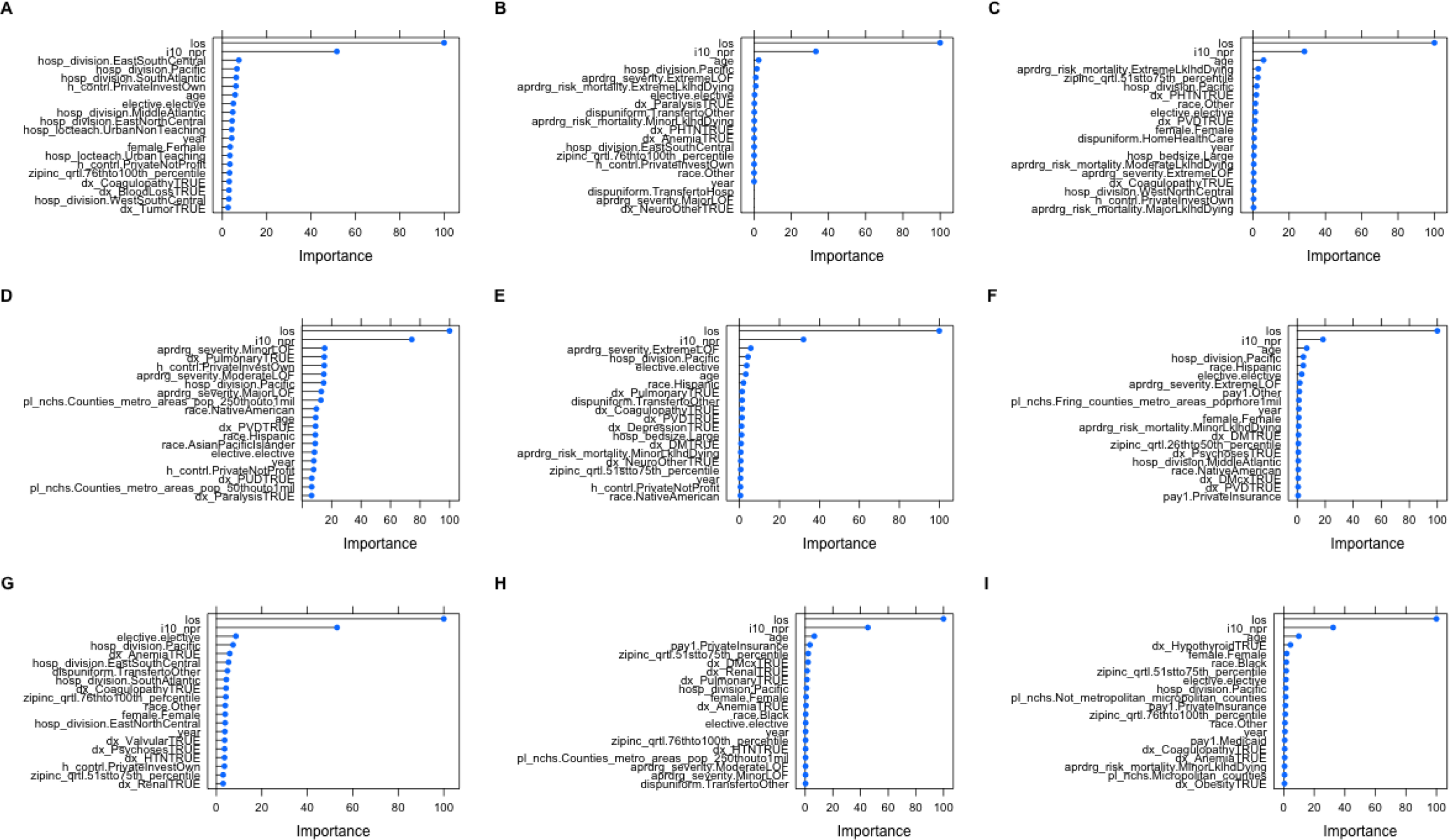
Variable Importance (VI) plots for each disease condition for the LM, GBM and XGB models, respectively. A-C: COPD exacerbation LM, GBM, XGB, respectively; D-E DKA episode LM, GBM, XGB, respectively; G-I: CHF exacerbation LM, GBM, XGB, respectively.

**Supplemental Figure 2.**
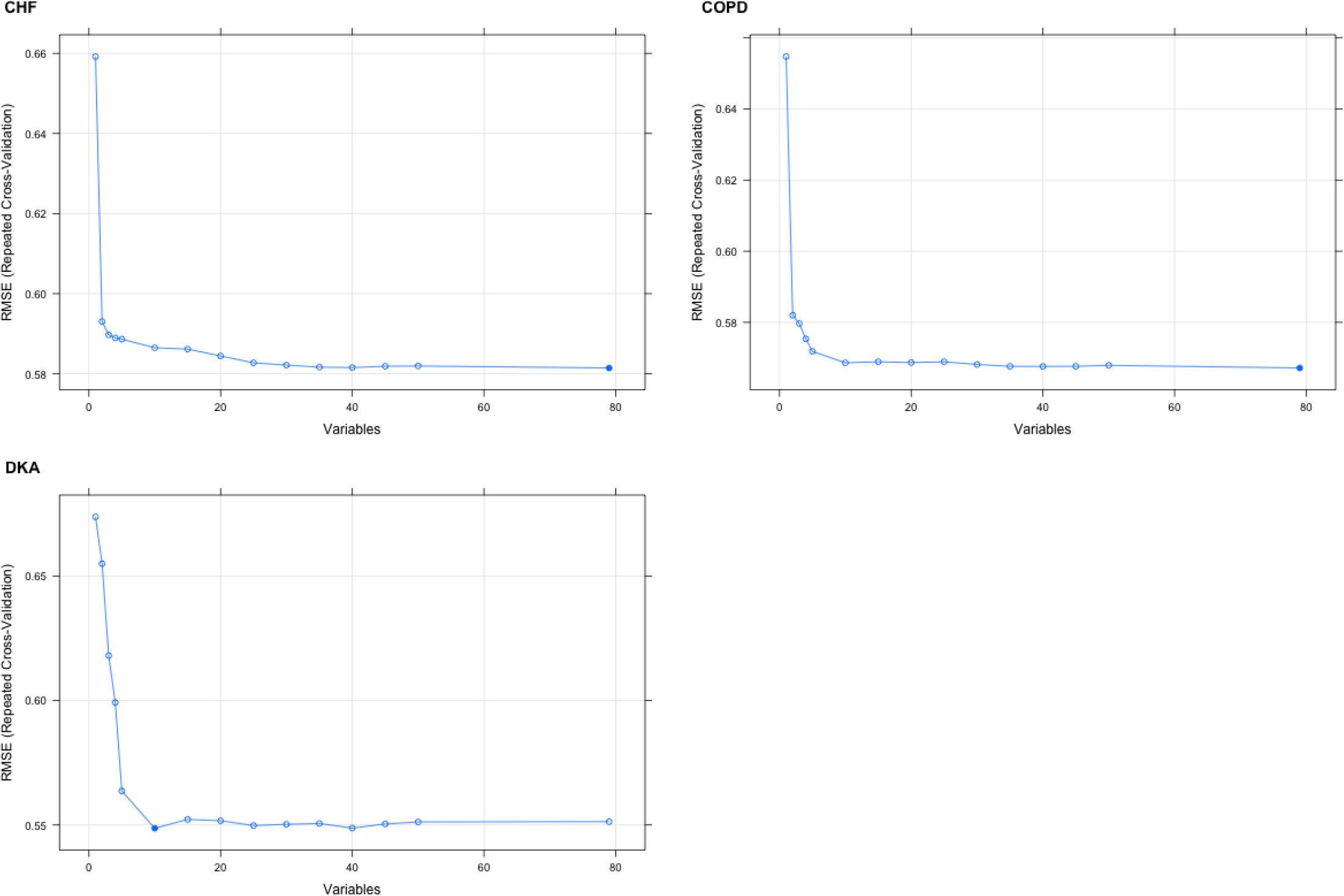
Optimal number of variables for each model determined using RFE algorithms

